# The content of biomedical papers predicts future clinical trials: a proof-of-concept framework tested on CAR-T cell therapy

**DOI:** 10.64898/2026.10.10.26365160

**Authors:** John Brennan, Justin Slater, Yan Yan

## Abstract

This paper shows that it is possible to predict which newly published biomedical research papers will be connected to subsequent clinical trials. We use CAR-T cell therapy as a proof-of-concept setting because its development can be followed from published research through clinical testing, regulatory approval, and commercialization. We reconstructed connections between research papers and clinical trials and trained machine-learning models to predict which papers were associated with research that progressed to clinical testing.

Using 65 paper-inherent indicators, including scientific topic, language, biomedical orientation, and team structure, the models achieved a cross-validated AUC of 0.908. The resulting rankings were also practically informative. In a retrospective analysis using no information generated after 2016, 80% of the top 20 papers came from laboratories associated with one of 13 predefined major CAR-T clinical or commercial outcomes.

As an additional finding, first-year citations alone achieved an AUC of 0.797, while provenance indicators based on author, journal, and institutional reputation performed less strongly and added little to the paper-inherent model.

Our results show that whether biomedical research papers will be connected to subsequent clinical trials can be predicted from newly published papers with high discrimination in CAR-T. Our findings could help research organizations and pharmaceutical companies identify papers, laboratories, and therapeutic programs that warrant closer evaluation before clinical outcomes are known.

## 1 Introduction

### 1.1 Why clinical translation is worth predicting

Clinical translation is the process of turning discoveries from basic science or laboratory research into practical methods that improve patient care. Studies tracking promising preclinical findings over time have found that only roughly 5–10% reach licensed clinical use [1, 2]. Proposed explanations include limitations in experimental design, animal models, and reporting practices [3].

This low rate of translation matters because advancing a discovery toward clinical testing is costly. An Investigational New Drug (IND) application requires manufacturing data, toxicology, and a regulatory dossier. A Phase I study requires clinical sites, detailed protocols, safety monitoring, and participants willing to enroll. Even at this earliest clinical stage, the average cost of a Phase I trial has been estimated at approximately $1.4 million to $6.6 million per study [4]. The financial commitment may therefore reach millions of dollars before researchers know whether a therapy will succeed clinically.

Research leaders must choose which programs receive additional experiments, personnel, infrastructure, and funding. Pharmaceutical R&D teams face a similar problem when deciding which programs to advance. Better early selection would reduce resources spent on programs that fail, thus becoming an important component of R&D productivity [5, 6].

For these reasons, clinical translation is worth predicting. A crucial element is that predictions need to be estimated early enough to inform research leadership decisions. If predictions of success can be made close to the time of publication, research leaders could more quickly and accurately identify research that warrants further support and funding. Pharmaceutical scouts and business-development teams could use the predictions to prioritize laboratories, technologies, and programs for closer evaluation. Previous studies have proposed that predictions could be used for research-portfolio evaluation [7, 8]. Such predictions would not replace scientific judgment or research management expertise. Their value would be in helping determine where limited time, expertise, and resources should be concentrated for more efficient research allocation.

### 1.2 The question

Can we predict which newly published biomedical research papers will be connected to subsequent clinical trials? In this study, a paper is considered connected when it has at least one direct clinical-trial connection or, when the evidence is indirect, connections to at least two separate clinical trials, as described in Section 3.3.

### 1.3 The claim

It is possible to predict which newly published biomedical research papers will be connected to subsequent clinical trials. We test whether that prediction can be made from three different kinds of information:

- The paper itself: what the research studies, how it is written, and how the research team is structured.
- Citations: how much attention the paper receives after it is published.
- Provenance: the reputation of the journal, institution, and authors behind the paper.

Our main finding is that the paper itself provides the strongest prediction. Paper-inherent indicators reach an AUC of 0.908. First-year citations reach 0.797. Journal and institutional prestige contribute almost no predictive signal. The important result is that we can predict which papers will be connected to subsequent clinical trials using information available in the paper at publication.

### 1.4 Contributions

This paper contributes to biomedical scientometrics by introducing new data, methods, and evidence for predicting which biomedical research papers will be connected to subsequent clinical trials.

This paper makes five contributions:

1. A labeled research dataset. We construct a dataset of 1,350 biomedical papers labeled according to whether they were connected to subsequent clinical trials.
2. A reproducible linking pipeline. We develop a public-data pipeline that connects clinical trials back to the research papers that preceded them.
3. Three predictive models. We train logistic regression, random forest, and gradient boosting models that rank newly published papers by their likelihood of being connected to subsequent clinical trials.
4. Evidence that the content of research papers is highly predictive. Indicators derived from the scientific content and structure of the paper reach an AUC of 0.908.
5. Evidence that the approach works beyond a single test. We evaluate the rankings against major CAR-T clinical and commercial outcomes and apply the same framework to checkpoint inhibitors to test how well it transfers to a second therapeutic area.

### 1.5 Overview of the approach

This study has three stages.

1. Build the data. We link CAR-T clinical trials back to the research papers that preceded them. This gives us papers that were connected to clinical trials and papers that were not.
2. Make the predictions. We use machine learning to estimate which newly published papers are most likely to be connected to a subsequent clinical trial. We train three different types of models: logistic regression, random forest, and gradient boosting.
3. Evaluate the predictions. We examine how well the models distinguish papers connected to subsequent clinical trials from those with no identified connection, compare the resulting rankings with major CAR-T clinical and commercial outcomes, and test the framework in a second therapeutic area.

## 2 Background

A therapeutic area is a field of medicine organized around a disease, condition, or type of treatment. CAR-T cell therapy area is the test case used in this paper. CAR-T is useful for this study because its development can be followed from published research, through clinical trials, to approved treatments and commercial outcomes.

### 2.1 What is CAR-T?

CAR-T cell therapy is a form of cancer treatment that modifies a patient’s own immune cells so they can recognize and attack cancer. T cells are immune cells that recognize and destroy abnormal or infected cells. Cancer can evade this response because tumor cells arise from the body’s own tissue and may not be effectively recognized by the immune system.

CAR-T therapy changes this recognition process. T cells are collected from a patient’s blood and genetically modified in the laboratory to express a chimeric antigen receptor, or CAR. The receptor is designed to bind a specific protein on the surface of cancer cells. In many B-cell cancers, that target is CD19. The modified T cells are then returned to the patient, where the CAR enables them to recognize and attack cells carrying the target protein [9].

CAR-T therapy is now an established treatment for several B-cell cancers, including certain leukemias and lymphomas [9]. The first CAR-T therapy was approved by the U.S. Food and Drug Administration in 2017 [10].

### 2.2 Why CAR-T is a strong test case

To test whether newly published papers can be used to predict connections to future clinical trials, we need a field with a clear historical record. The research must be old enough for trials and commercial outcomes to have already occurred, but recent enough that the papers, trials, and laboratories can still be linked reliably.

CAR-T fits these requirements well. Key papers were published in 2015 and 2016, and clinical development followed quickly. By the end of 2016, 113 CAR-T trials were registered on ClinicalTrials.gov. The first FDA-approved CAR-T products arrived in 2017, followed by additional approvals and major commercial activity over the next several years.

The outcomes are well documented. Between 2017 and 2022, the field produced six FDA-approved products, two acquisitions valued above $1 billion, and five major company foundings or public-market events. These events can be traced back to specific laboratories and institutions using FDA records, company announcements, and securities filings. The 13 outcomes are listed in Section 6.1 and used as the retrospective benchmark.

This gives us a field in which we can make predictions using only information available at an earlier point in time and then compare those predictions with outcomes that are already known.

### 2.3 Prior work

Previous studies have shown that characteristics of biomedical papers can help to predict what happens with the research afterwards. For example, studies have used characteristics of papers to predict outcomes such as connections to clinical research, scientific impact, and other forms of translational impact.

Hutchins et al. [8] is a close predecessor to our study. They built a model to predict whether a biomedical research paper would become connected to clinical research. Their predictors included citation rate, characteristics of the paper’s citing network, and MeSH-based measures of biomedical content. One of their content-based predictors was the Triangle of Biomedicine, introduced by Weber [7], which classifies papers according to whether their subject matter is more focused on humans, animals, or molecules and cells. Using two years of post-publication data, their model achieved an AUC of 0.80, with citation activity and characteristics of the citing network among the strongest predictors. Although it is intuitive that human-oriented papers are going to be closer to clinical trials than molecular- or animal-oriented studies, the Triangle of Biomedicine was helpful because we used it as one of the paper-inherent indicators in our own prediction framework.

Other studies have examined related prediction problems. Wang et al. [11] modeled the long-term citation trajectories of scientific papers and showed that early citation patterns contain information about future scientific impact. Nelson et al. [12] used deep-learning models of title and abstract content to forecast downstream translational impact, indexed by inclusion in patents, guidelines, or policy documents, and reported that content-based models outperformed citations and metadata alone. Wang et al. [13] approached the problem from the opposite direction of this study, using information from registered clinical trials to predict whether those trials would later produce publications. More broadly, Fortunato et al. [14] reviewed the growing science-of-science literature on predicting and understanding scientific outcomes.

This body of research shows that characteristics of biomedical publications can predict several downstream scientific and translational outcomes. Most of the studies don’t provide information that would help us with a comparative AUC score. Our focus is on how newly published papers can be used to predict which papers will be connected to subsequent clinical trials. We examine this question in CAR-T cell therapy and also consider citations and provenance as additional sources of predictive information.

## 3 Data

To build the prediction models, we needed to identify which research papers were connected to clinical trials. No existing database provides a complete record of these connections, so we constructed the dataset by combining multiple sources of evidence.

Some trials directly identify the earlier research papers connected to them, but many do not. We combined several sources to recover these missing paper-to-trial connections. When a paper was directly linked to a trial, we treated that as strong evidence. When the connection was less direct, we required additional supporting evidence before labeling the paper as positive.

### 3.1 Clinical-trial corpus

We began with all 596 CAR-T clinical trials registered on ClinicalTrials.gov [15] between 2017 and 2021 that included a Phase I or Phase II component.

Phase I trials are the earliest studies in humans and focus primarily on safety, side effects, and dose. Phase II trials begin to evaluate if the treatment produces the intended effect while continuing to monitor safety [16]. We focus on these phases because they provide clear evidence that a treatment has entered human testing.

ClinicalTrials.gov allows trials to have more than one phase designation, so the categories overlap:

- 524 trials include Phase I;
- 239 trials include Phase II;
- 167 trials are designated Phase I/II; and
- 7 trials are designated Phase II/III.

A named lead investigator was available for 446 of the 596 trials, providing another source of information for connecting trials to earlier research.

### 3.2 Reconstructing paper-to-trial connections

We reconstructed the research history behind these trials using ClinicalTrials.gov [15], PubMed, PubMed Knowledge Graph 2.0 [17], and OpenAlex [18].

We began with the strongest form of evidence: an explicit reference connecting an earlier paper to a clinical trial. ClinicalTrials.gov sometimes records these papers directly, and PKG 2.0 provides additional structured paper-to-trial connections [17].

Because these direct references are incomplete, we used additional methods to recover missing connections. These included matching trial identifiers to publications, matching named trial investigators to their earlier CAR-T research, and tracing citations backward from papers reporting trial results.

The last method was particularly useful. A paper reporting the results of a trial usually cited earlier research related to the treatment being tested. We identified those cited papers and retained only papers published before the trial was registered.

This procedure recovered 21,512 candidate papers across 542 of the 596 trials, providing coverage for 90.9% of the trial corpus.

Not all of these connections provided equally strong evidence. We therefore distinguished between two levels:

- Direct evidence: an earlier paper was explicitly connected to a clinical trial.
- Indirect evidence: an earlier paper was cited by a paper reporting the results of a clinical trial.

Direct connections were accepted as strong evidence. Indirect connections were subjected to an additional requirement before they could be used as positive examples.

### 3.3 Constructing the final labels

We restricted the analysis to papers published in 2015 and 2016, the period used for prediction. For papers supported by direct evidence, one trial connection was sufficient. For papers supported only by indirect evidence, we required connections to at least two separate clinical trials. This rule reduced the likelihood that a paper was labeled positive merely because it was commonly cited as background. Repeated connections across independent trials provided stronger evidence that the paper was meaningfully connected to subsequent clinical testing.

Figure 1 summarizes the two-branch label-construction pipeline described in this section.

**Fig. 1.**
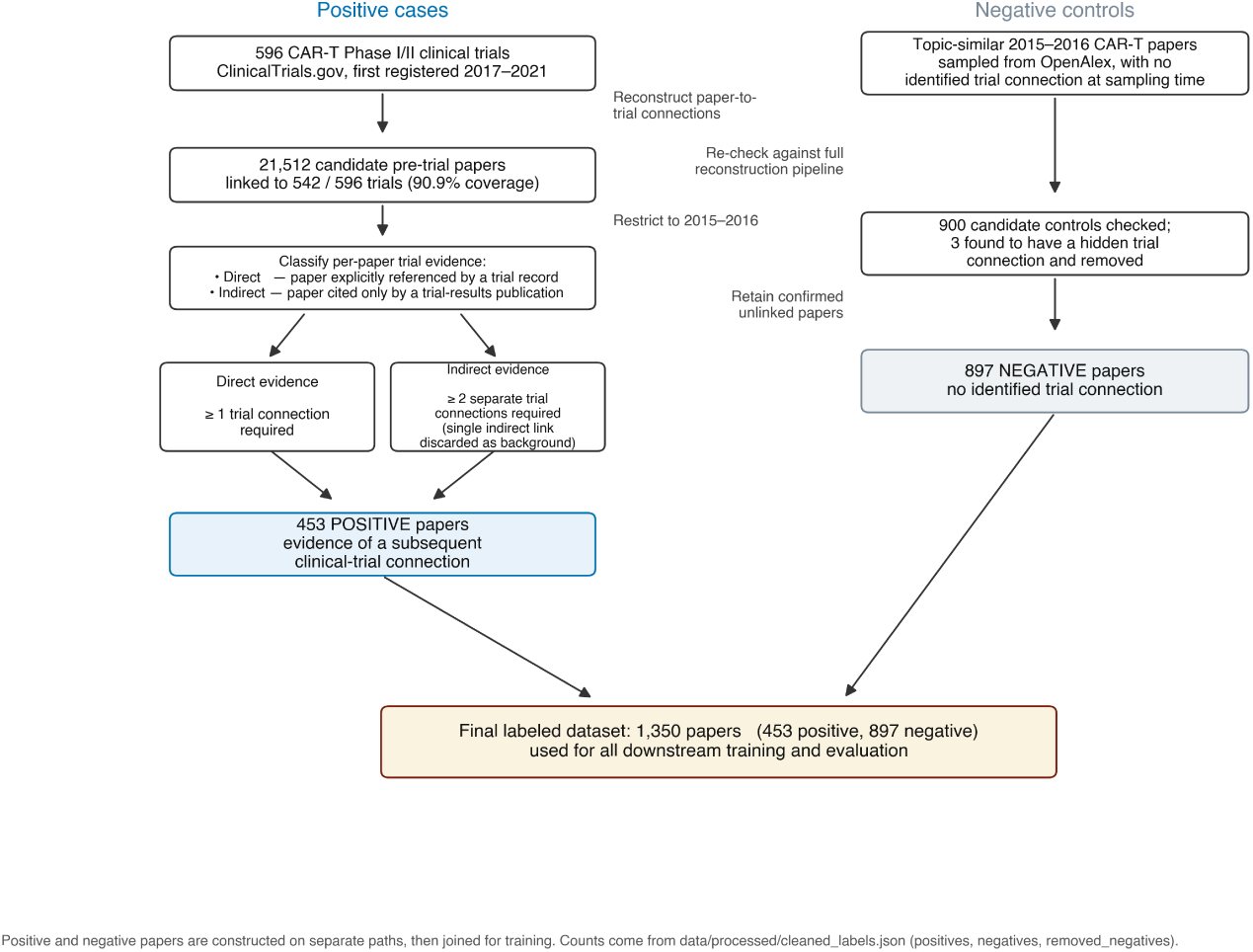
Case-control construction of the 1,350-paper labeled corpus. Positives are 2015–2016 papers linked to a CAR-T Phase I/II trial by direct reference (*≥* 1 link) or backward citation from a trial-results paper (*≥* 2 links). Negatives are topic-similar 2015–2016 CAR-T papers with no identified trial link after re-checking.

Applying these criteria produced 453 positive papers. We also assembled 897 negative papers for which no clinical-trial connection was identified. These papers were rechecked against the same sources used in the reconstruction pipeline to reduce missed connections.

The final dataset therefore contains 1,350 papers: 453 with evidence of a later clinical-trial connection and 897 with no identified connection. For papers with an identified trial connection where the lag could be measured, the median time from publication to the first connected trial record was two years (Figure 2).

**Fig. 2.**
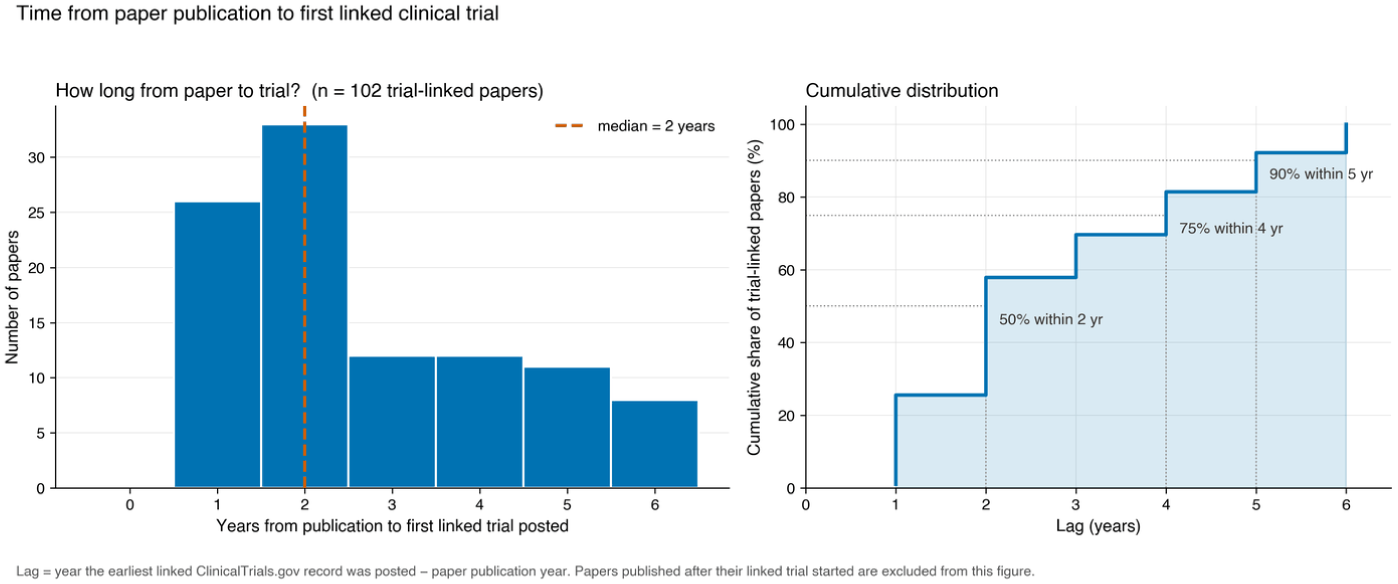
Time from publication to the first connected ClinicalTrials.gov record, for papers with an identified trial connection where the lag could be measured (*n* = 102). Papers published after their connected trial began are excluded.

These labels represent evidence that a paper was connected to later clinical testing. They do not imply that every positive paper directly caused or motivated a specific trial.

## 4 Indicators and Models

With our constructed dataset, we next describe each paper using a set of measurable indicators. These indicators capture different kinds of information about the research, the attention it receives, and the people and institutions behind it. We then use the indicators as inputs to three machine-learning models.

### 4.1 Building the indicator set

We developed the indicator set from three sources.

First, we drew from previous work. Hutchins et al. [8] showed that the biomedical orientation of a paper can help predict if it becomes connected to clinical research. We therefore include the Triangle of Biomedicine measures introduced by Weber [7], in the form adapted by Hutchins et al. [8].

Second, we created indicators describing characteristics that can be observed directly from a paper. These include subject matter, language, disease references, author count, institution count, and other characteristics of the research and research team.

Third, we added external signals commonly used to assess research. These include citations, journal prestige, institutional standing, and author reputation. Including these measures allows us to compare information contained in the paper with information surrounding it.

Together, these sources produced 77 indicators.

### 4.2 The three indicator families

We divide the 77 indicators into three families.

Paper-inherent indicators (65). These describe the research itself and can be derived from the paper.

- Paper and team properties (11). Author count, number of institutions, abstract length, review-article status, large-team status, and indicators for mentions of myeloma, lymphoma, CD19, first-in-human language, clinical concepts, and clinical trials.
- Triangle of Biomedicine indicators (4). These measure how much of a paper’s subject matter concerns humans, animals, or molecules and cells using its Medical Subject Headings (MeSH) terms [19]. We also include the total number of MeSH terms. For example, if two of eight MeSH terms are classified as human-related, the human fraction is 0.25. These measures follow Weber [7] and Hutchins et al. [8].
- Semantic indicators (50). We use the title and abstract of each paper to capture differences in topic and language. TF-IDF converts words and two-word phrases into numerical values based on how informative they are across the corpus. This initially produces roughly 20,000 text dimensions. We then reduce these to 50 dimensions using truncated singular value decomposition. The resulting indicators summarize patterns in what papers discuss and how they describe the research.

Citation indicators (2). These measure citations accumulated during the first full year after publication. We include the raw citation count and its logarithmic transformation.

Provenance indicators (10). These describe the reputation of the authors, journal, and institutions associated with the paper. They include first- and last-author h-index and publication count, transformed versions of those measures, journal prestige, and institutional standing. Some author-reputation measures reflect current rather than historically reconstructed values, which should be considered when interpreting their predictive contribution.

### 4.3 Models and evaluation

We train three types of machine-learning models using the same labeled papers and indicator sets:

- **Logistic regression** assigns a weight to each indicator and combines those weights to estimate the probability that a paper will be connected to a clinical trial. Its main advantage is that the estimated weights can be examined directly.
- **Random forest** builds many decision trees and combines their predictions. Each tree repeatedly divides papers into groups based on their indicator values.
- **Gradient boosting** (specifically HistGradientBoosting, HistGBM) also uses decision trees, but builds them sequentially. Each new tree focuses on correcting errors made by the trees built before it.

All three models, the TF-IDF and truncated singular value decomposition steps, and the cross-validation procedure are implemented in scikit-learn [20].

We use three model types because they capture relationships between the indicators and paper-trial connections in different ways. If they produce similar results, this gives us greater confidence that the findings are not specific to one modeling approach.

We evaluate the models using five-fold cross-validation. The 1,350 papers are divided into five groups. Each model is trained on four groups and tested on the fifth. This process is repeated five times so that every paper appears in a test group once, and performance is then averaged across the five tests.

Unless otherwise stated, the reported AUC for each indicator set is the average AUC across logistic regression, random forest, and gradient boosting.

We measure performance using the Area Under the receiver operating characteristic Curve, or AUC. AUC measures how well the model distinguishes papers that were connected to clinical trials from papers that were not. An AUC of 0.50 indicates performance no better than chance, while an AUC of 1.00 indicates perfect separation between the two groups.

## 5 Results

### Main result: paper-inherent indicators strongly predict which papers are connected to subsequent clinical trials

The main question in this study is whether it is possible to predict which newly published biomedical research papers will be connected to subsequent clinical trials. Using only the 65 paper-inherent indicators, the models achieve an AUC of 0.908 *±* 0.017. An AUC above 0.90 is generally considered excellent discrimination [21]. In practical terms, this means that if the model is given one paper that was later connected to a clinical trial and one that was not, it will usually assign the higher score to the connected paper.

The main result is that we can predict whether a research paper will be connected to a subsequent clinical trial with high accuracy. Moreover, this strong prediction can be made using information contained in the research paper itself. We next examine what kinds of information within the paper contribute to this performance and whether adding citations or measures of reputation improves the prediction.

### 5.1 What the paper-inherent indicators capture

The 65 paper-inherent indicators describe several different aspects of the research.

The Triangle of Biomedicine indicators describe whether the paper is focused more heavily on humans, animals, or molecular and cellular research (Figure 3).

**Fig. 3.**
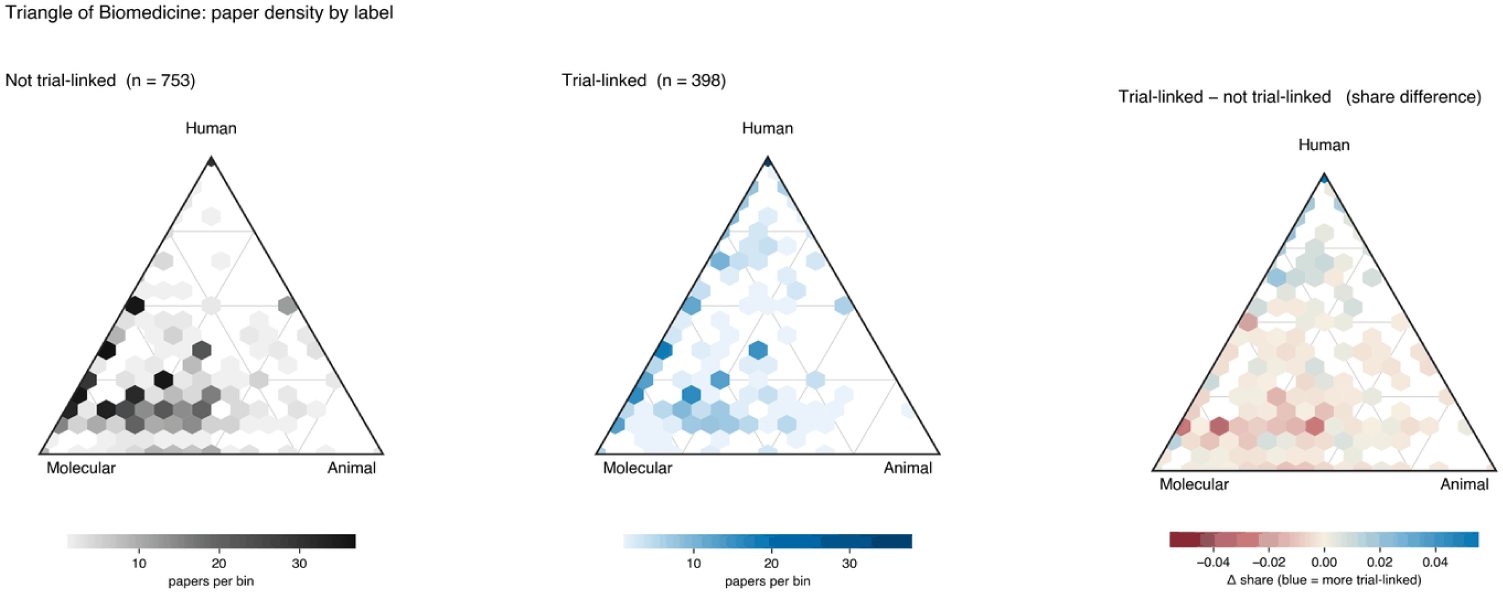
Triangle of Biomedicine positions for papers with and without an identified trial connection. Each hexagon is binned by the human, animal, and molecular fractions of a paper’s MeSH terms. The right panel shows the difference in share between the two groups. Panels include only papers with usable MeSH terms (*n* = 1,151).

The semantic indicators capture patterns in the title and abstract. They allow the models to distinguish papers based on differences in topic and language that are more detailed than the broader MeSH (Medical Subject Headings) categories (Figures 4, 5, and 6).

**Fig. 4.**
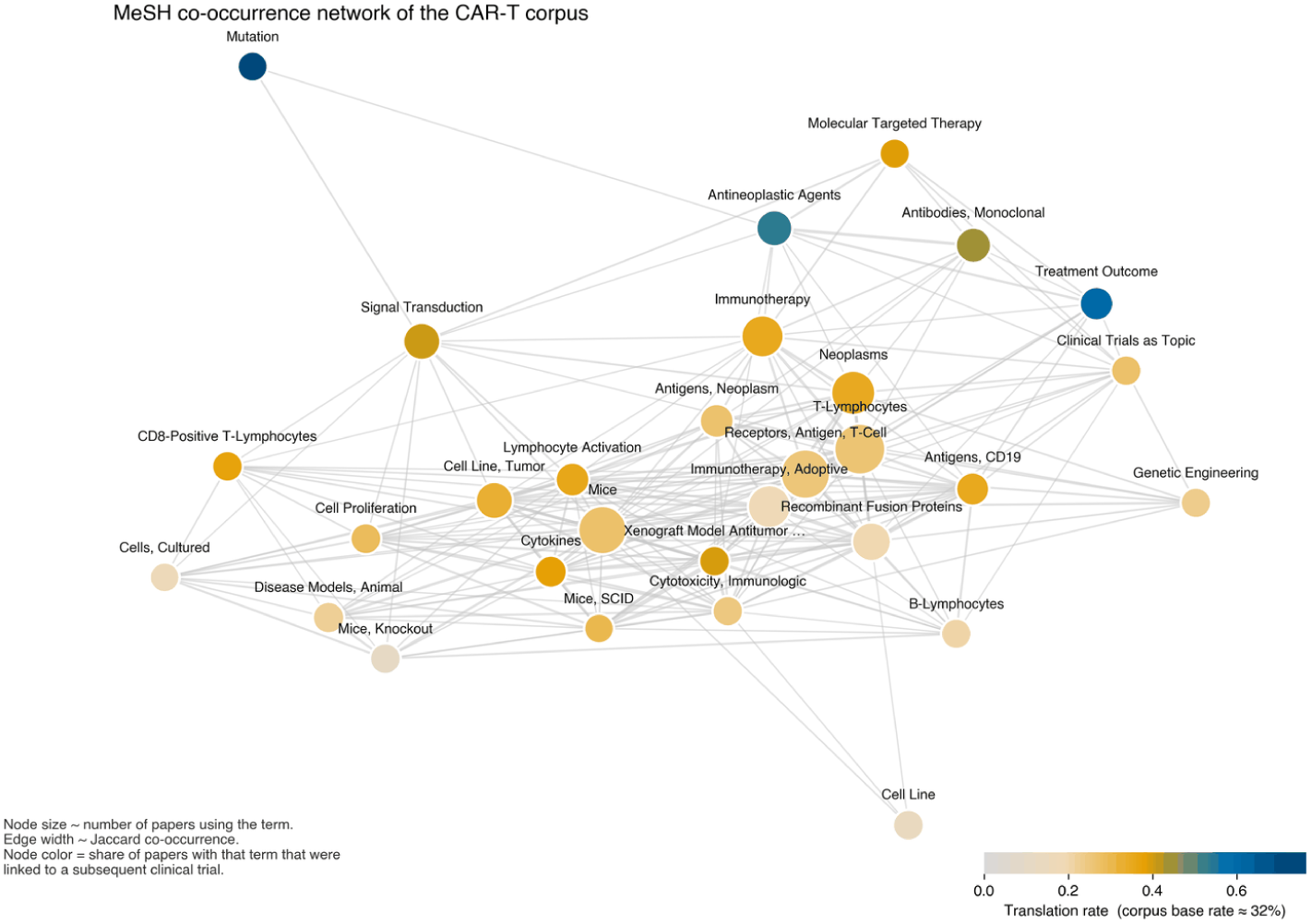
MeSH co-occurrence network for the CAR-T corpus. Node size reflects the number of papers using the term, edge width reflects Jaccard co-occurrence, and node colour reflects the share of papers using that term that were connected to a subsequent clinical trial.

**Fig. 5.**
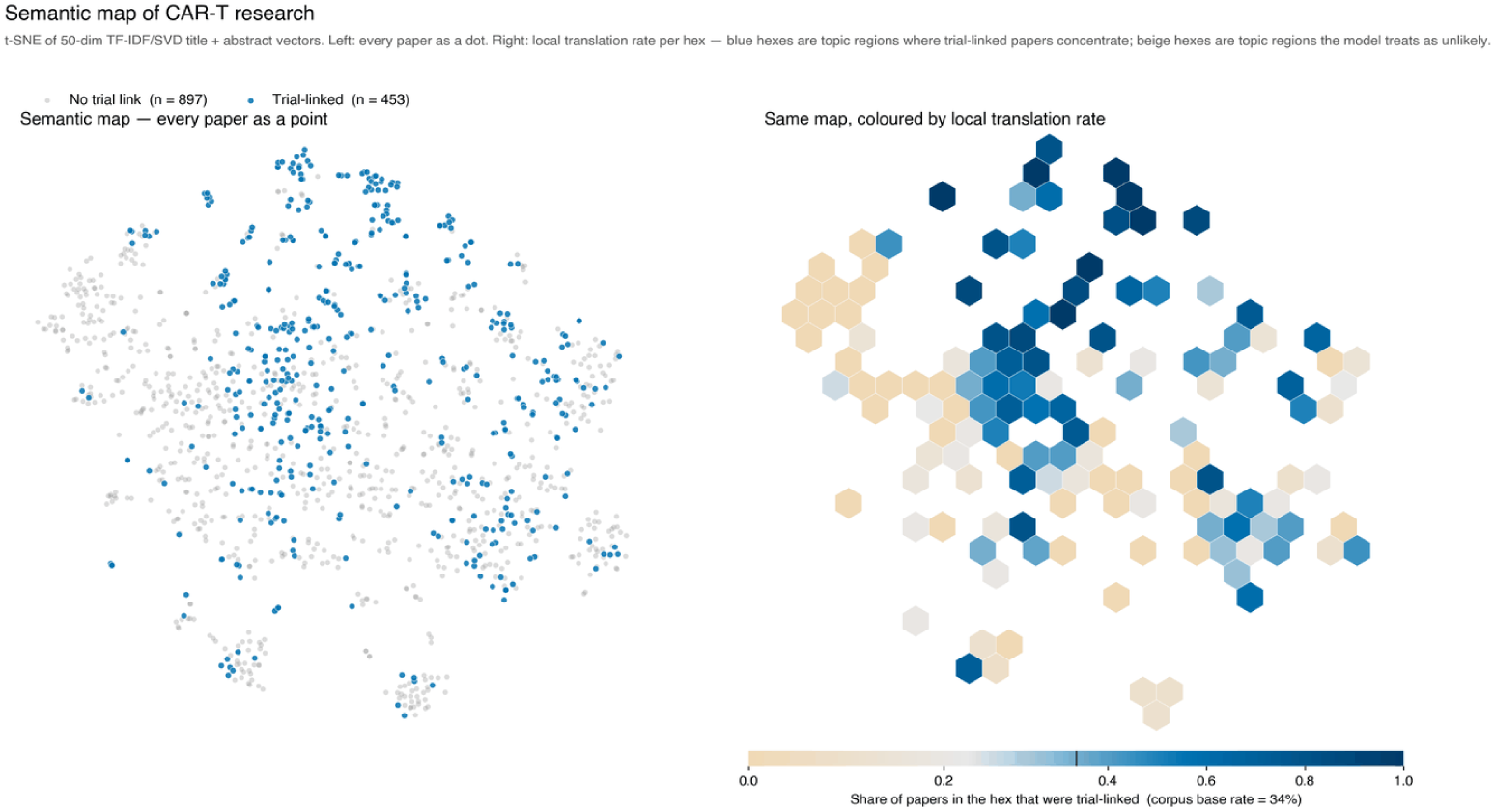
Semantic map of the CAR-T corpus. Left: t-SNE projection of the 50-dimensional TF-IDF/SVD title and abstract vectors, with one point per paper. Right: the same projection with each hexagonal region coloured by the share of its papers with an identified trial connection, against a corpus base rate of 34%.

The paper and team indicators capture additional characteristics such as team size, number of institutions, review status, and references to specific diseases or clinical concepts (Figure 7).

**Fig. 6.**
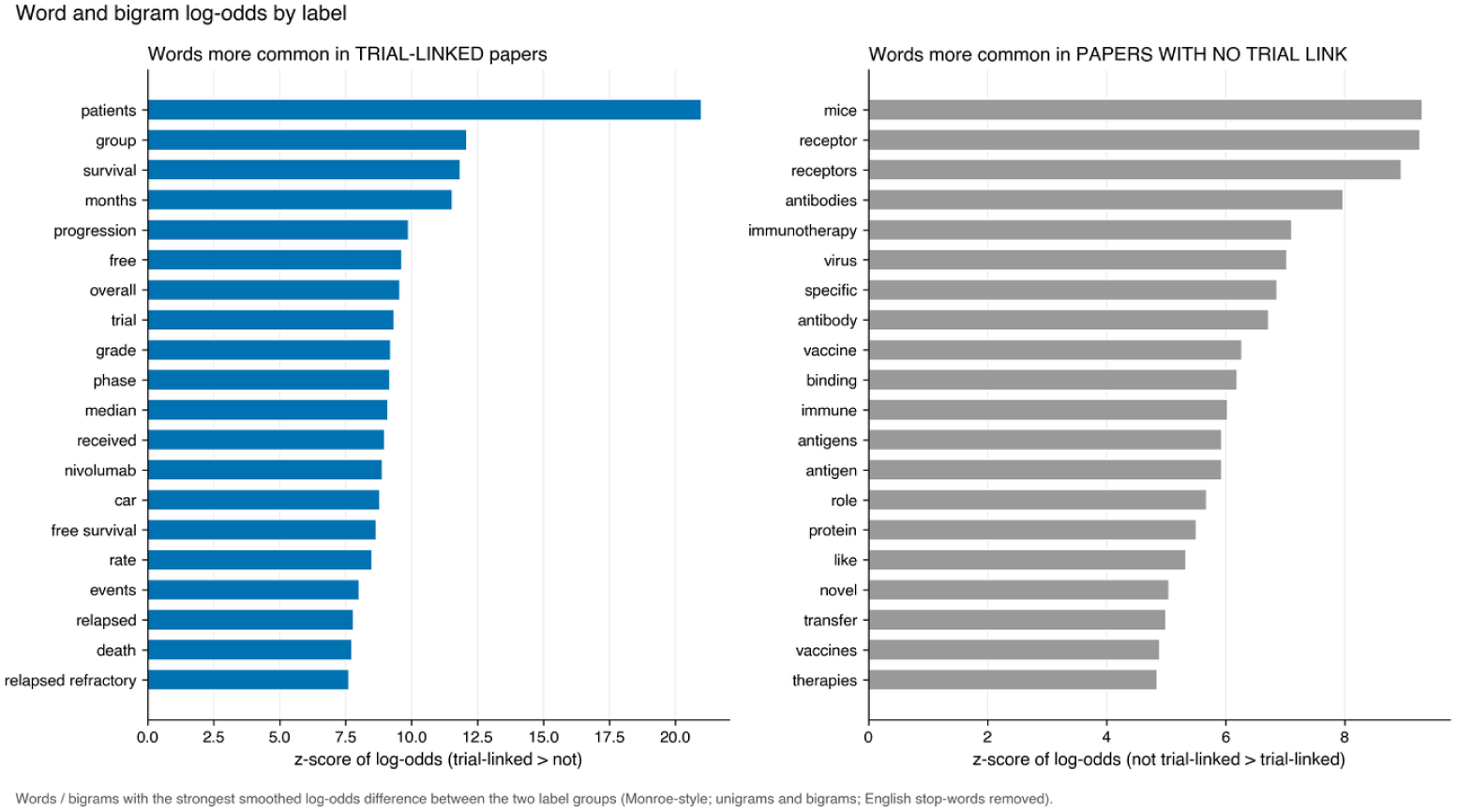
Words and two-word phrases with the largest smoothed log-odds difference between the two label groups. Unigrams and bigrams, with English stop-words removed.

**Fig. 7.**
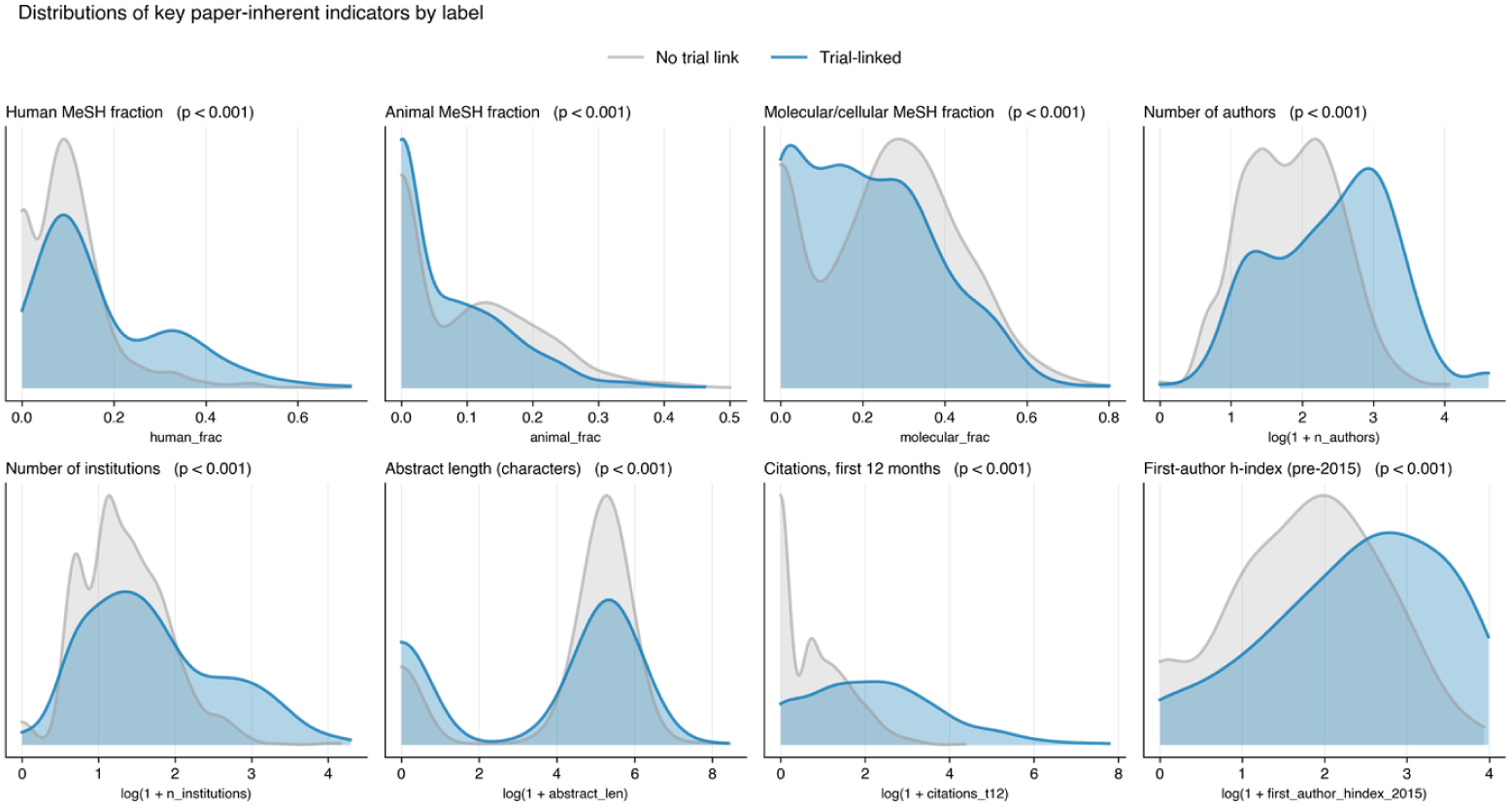
Distributions of eight paper-inherent indicators by label. Values for count-based indicators are shown on a logarithmic scale.

Together, these indicators describe the subject matter, language, and structure of the paper (Figure 8).

**Fig. 8.**
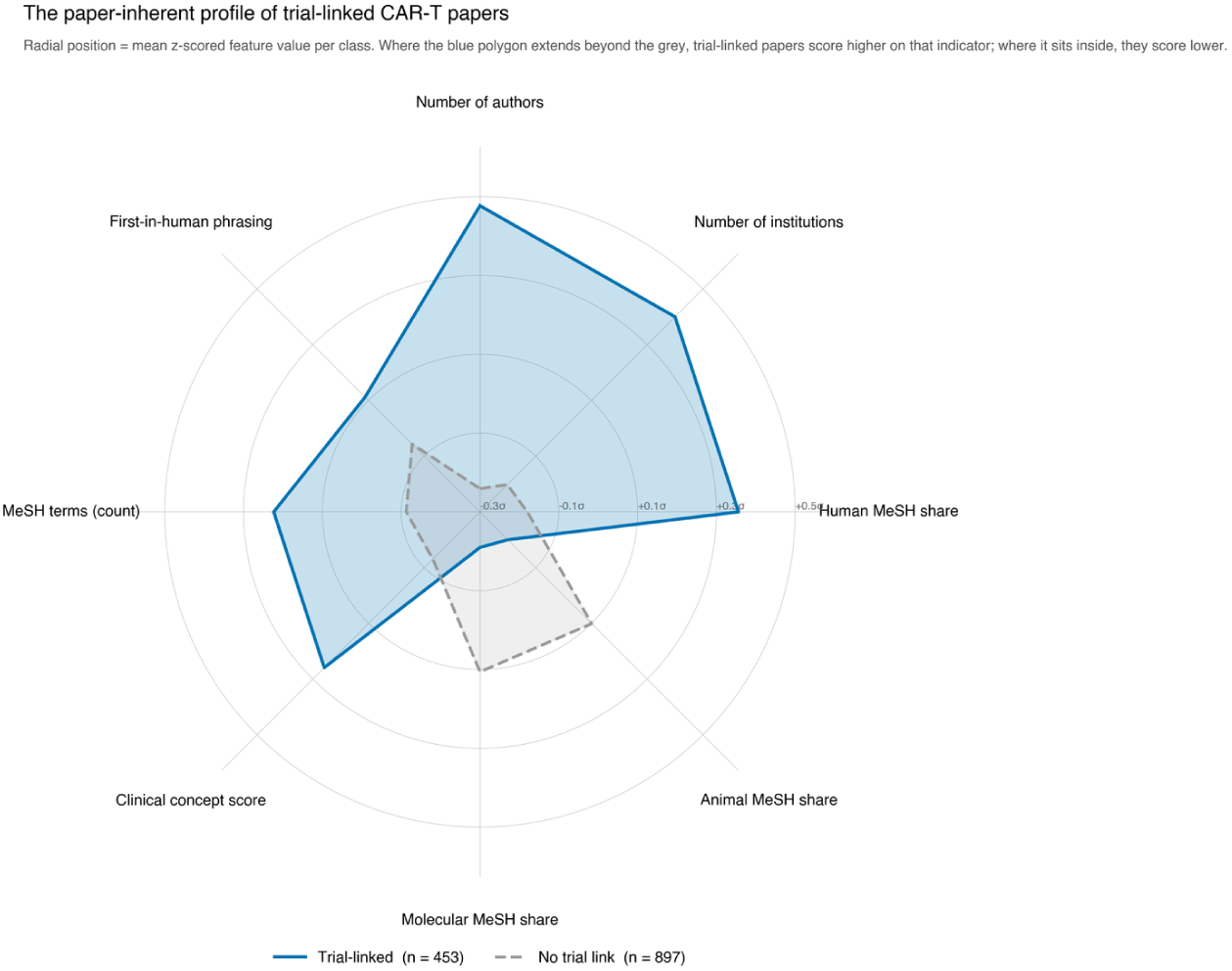
Mean *z*-scored values of eight paper-inherent indicators for each label group. Values are standardized across the full corpus, so each polygon shows where that group sits relative to the corpus mean.

### 5.2 Comparing paper information with citations and provenance

We tested if information outside the content of the paper improves the prediction. We compared four sets of indicators while keeping the same papers, cross-validation folds, and model settings (Tables 1 and 2, and Figure 9).

**Fig. 9.**
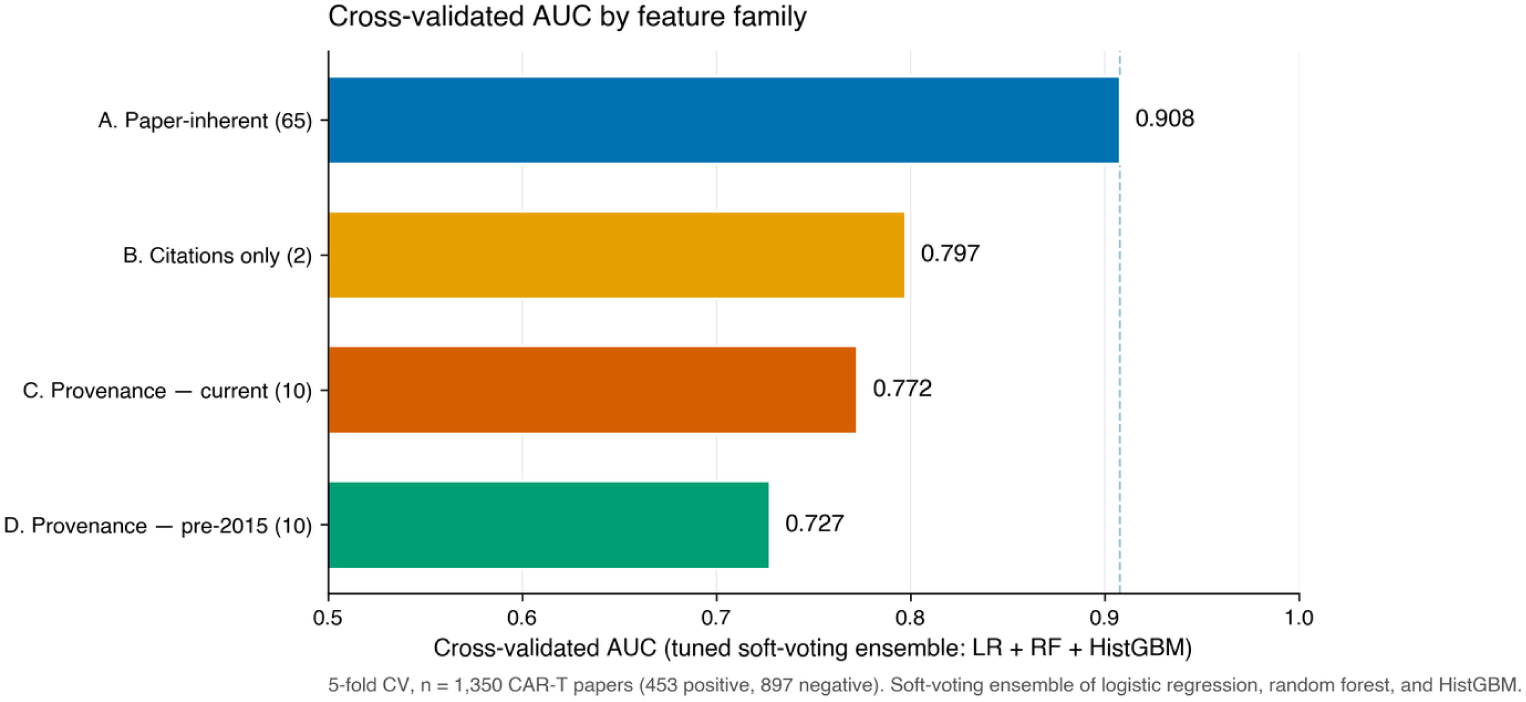
Cross-validated AUC for each indicator family evaluated on its own. The dashed line marks the paper-inherent result.

**Table 1.** Cross-validated AUC and PR-AUC by indicator set, built up from the paper-inherent baseline.

| Model | Indicators | CV AUC | CV PR-AUC | $\Delta$ AUC |
| --- | --- | --- | --- | --- |
| Paper-inherent only | 65 | $0.908 \pm 0.017$ | $0.868 \pm 0.017$ | — |
| + provenance (current) | 75 | $0.912 \pm 0.013$ | $0.872 \pm 0.020$ | +0.0041 |
| + provenance (pre-2015) | 75 | 0.907 | $0.868 \pm 0.014$ | −0.0011 |
| + citations | 67 | $0.923 \pm 0.015$ | $0.891 \pm 0.014$ | +0.0149 |
| + citations + provenance (current) | 77 | 0.923 | $0.892 \pm 0.016$ | +0.0150 |
| + citations + provenance (pre-2015) | 77 | $0.923 \pm 0.013$ | $0.894 \pm 0.014$ | +0.0155 |

Note: All models use the same papers, cross-validation folds, and model settings. $\Delta$ AUC is measured against the paper-inherent baseline of 0.908. Baseline PR-AUC ≈ 0.336 (corpus base rate).

**Table 2.**
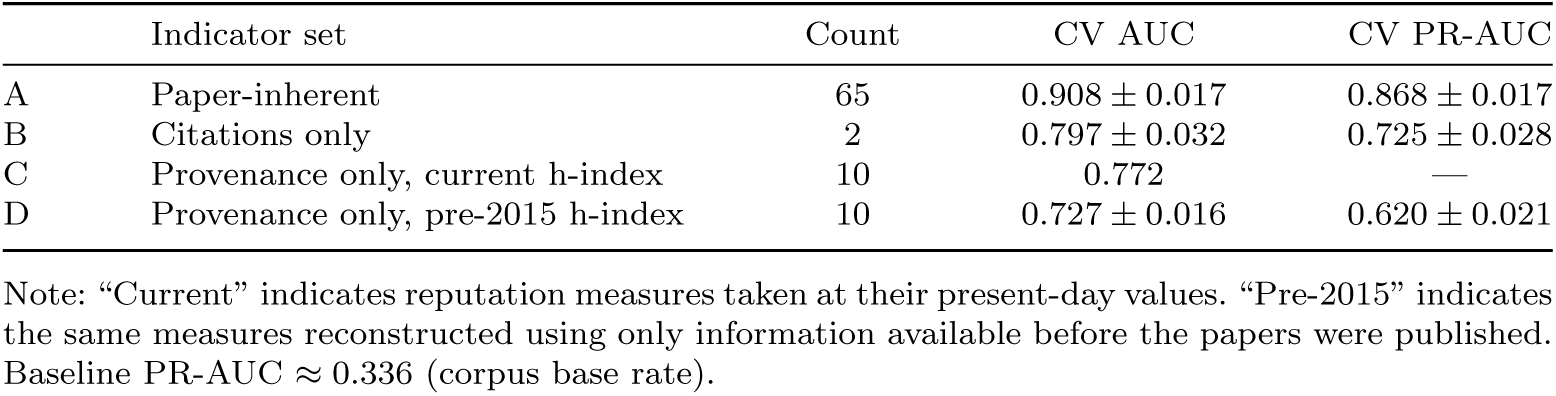
Each indicator family evaluated in isolation.

|  | Indicator set | Count | CV AUC | CV PR-AUC |
| --- | --- | --- | --- | --- |
| A | Paper-inherent | 65 | $0.908 \pm 0.017$ | $0.868 \pm 0.017$ |
| B | Citations only | 2 | $0.797 \pm 0.032$ | $0.725 \pm 0.028$ |
| C | Provenance only, current h-index | 10 | 0.772 | — |
| D | Provenance only, pre-2015 h-index | 10 | $0.727 \pm 0.016$ | $0.620 \pm 0.021$ |

Note: “Current” indicates reputation measures taken at their present-day values. “Pre-2015” indicates the same measures reconstructed using only information available before the papers were published. Baseline PR-AUC ≈ 0.336 (corpus base rate).

The final dataset is moderately imbalanced (453 positive of 1,350 papers, 33.6%). We report cross-validated AUC as the headline discrimination measure, but note that AUC can overstate practical usefulness under class imbalance; we therefore report PR-AUC alongside AUC in Tables 1 and 2 and revisit the imbalance issue in Section 9 [22].

Adding provenance indicators to the paper-inherent model increases AUC only slightly, from 0.908 to 0.912. Adding first-year citations increases performance further, producing a full-model AUC of 0.923. First-year citations are also predictive, with an AUC of 0.797, although their performance is notably lower than that of the paper-inherent indicators.

This comparison is especially important because citations require time to accumulate. The citation indicators use a full year of information after publication, while the paper-inherent indicators are derived from the research paper itself. The results therefore show that strong predictive information is already present in the paper. Citations improve the prediction, but they are not necessary to achieve high performance (Figure 10).

**Fig. 10.**
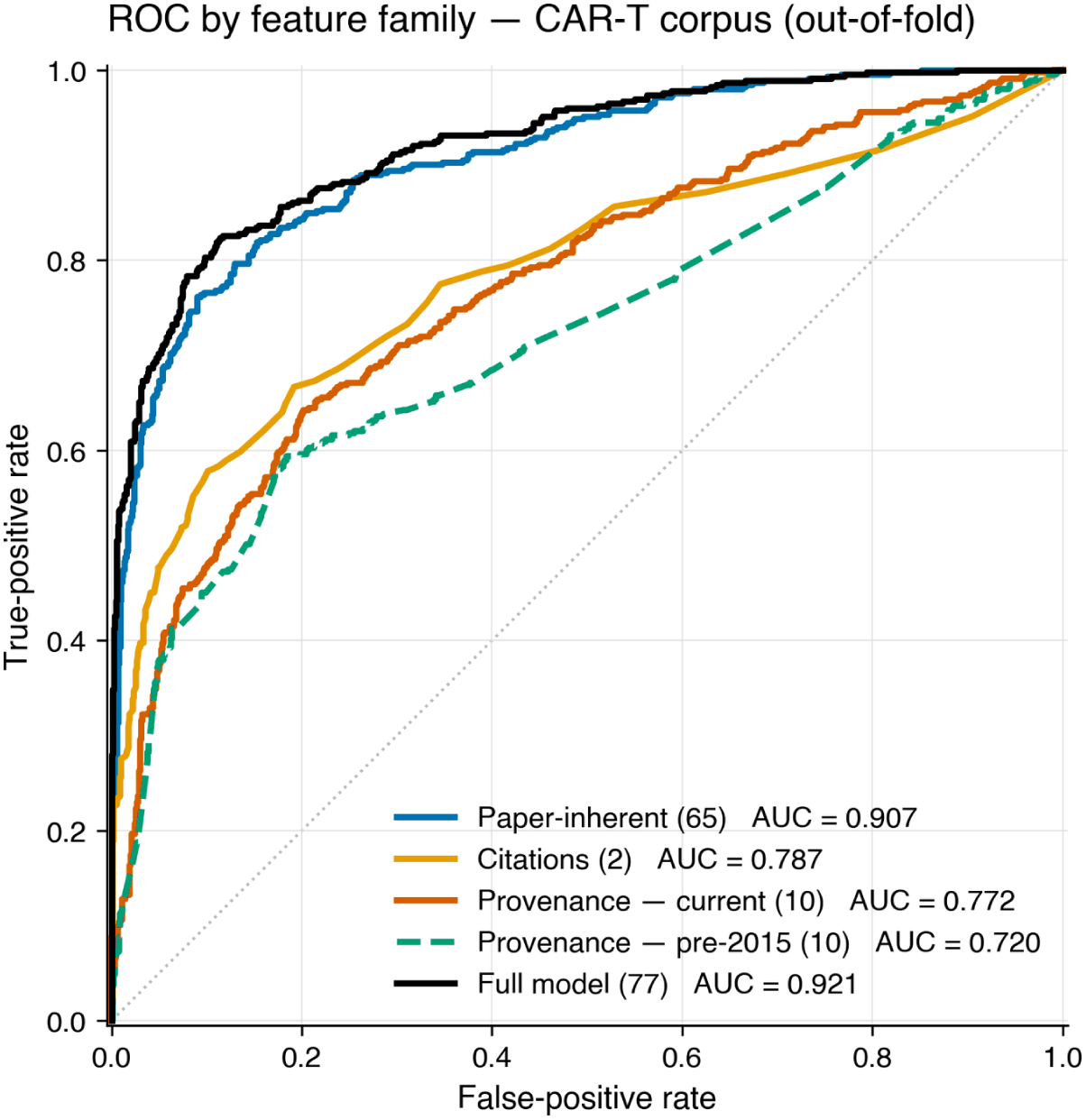
Out-of-fold ROC (Receiver Operating Characteristic Curve) curves by indicator family. AUC values in the legend pool all held-out predictions into a single curve and therefore differ slightly from the cross-validated means reported in Table 1 and Figure 9.

### 5.3 How much does provenance add?

We also examined whether the paper-inherent indicators might simply be identifying prestigious researchers or institutions.

One possible explanation is that papers from well-known laboratories are more likely to be connected to clinical trials because those laboratories have greater funding, clinical infrastructure, or industry connections. If this were the main reason for the model’s performance, measures of author, journal, and institutional reputation should add substantial predictive information.

Adding all 10 provenance indicators to the paper-inherent model increases AUC from 0.908 to 0.912 (Figure 11).

**Fig. 11.**
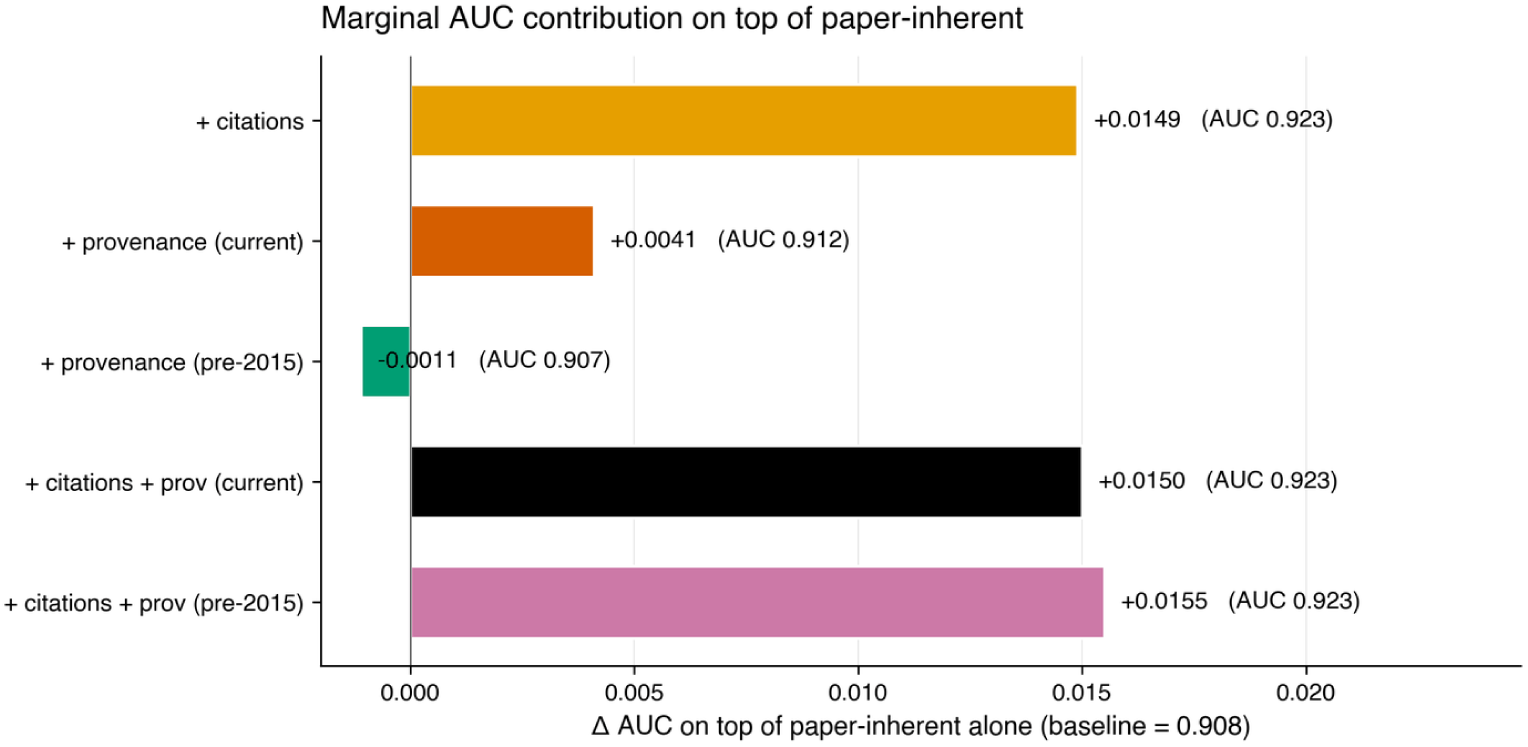
Change in cross-validated AUC when each external indicator family is added to the paper-inherent model, measured against a baseline of 0.908.

Individual indicators for elite institutions and top-ranked journals also receive little weight across the models. For example, the elite-institution indicator has a standardized logistic-regression coefficient of *−*0.054 and low importance in both the random-forest and gradient-boosting models. The top-journal indicator shows a similarly small contribution.

These results suggest that, within the CAR-T papers in this study, knowing who wrote the paper, where they work, and where the paper was published adds relatively little once the information contained in the paper itself is known.

This result should be interpreted cautiously. The subject of provenance is vast and multi-factoral. CAR-T research is concentrated among major research institutions, so there may be limited variation in institutional prestige within the dataset. In addition, some author-reputation measures use current rather than historical values, as discussed in Section 4.2.

## 6 Could the Prediction Framework Help Identify Promising Laboratories Early?

The results so far show that information contained in a research paper can be used to predict whether that paper will be connected to a subsequent clinical trial. We next ask whether these predictions could have been useful for pharmaceutical scouting and investment decisions.

Pharmaceutical companies increasingly look outside their own R&D organizations for new technologies and therapeutic programs, using partnerships, licensing, and acquisitions to access external innovation [23]. In 2016, CAR-T was a rapidly developing therapeutic field, creating a difficult screening problem for pharmaceutical scouts. Thousands of relevant papers were available, but it was not clear which research groups were working on programs with the strongest potential for clinical development.

The practical value of the prediction framework would therefore be to help identify which papers, laboratories, and research programs deserved the closest attention for possible investment, partnership, licensing, or acquisition.

To test this, we used the prediction framework to evaluate CAR-T papers published in 2015 and 2016 and identify which papers appeared most likely to be connected to subsequent clinical trials.

We then used those paper-level predictions to identify the laboratories behind the most promising research. The connection between the prediction framework and the commercial outcomes is:

- **Paper:** the framework evaluates a newly published CAR-T paper.
- **Clinical potential:** the framework gives a higher score to papers that appear more likely to be connected to a subsequent clinical trial.
- **Laboratory:** we identify the laboratory responsible for each highly scored paper, using the definition given below. In this work, a laboratory is a research group led by a principal investigator. We identify a laboratory by the name of its principal investigator and the institution where the group was based at the time of publication. The institution is included because the same investigator may lead a research group at different institutions over time. These are treated as distinct laboratory affiliations for attribution purposes.
- **Clinical or commercial outcome:** we check whether that laboratory is associated with a major CAR-T outcome, such as an FDA-approved product, a major acquisition, or a company founding or public-market event.

The framework therefore does not predict an acquisition, FDA approval, or company event directly. Instead, it identifies papers that appear most likely to be connected to subsequent clinical trials, and we test whether those papers came from laboratories that went on to achieve major clinical and commercial outcomes.

The question is therefore practical: if a pharmaceutical company had used the framework in 2016 to help decide which CAR-T research groups and programs were most worth investigating for investment, licensing, partnership, or acquisition, would the highest-scoring papers have directed attention toward laboratories associated with major clinical and commercial outcomes?

### 6.1 Defining the Clinical and Commercial Benchmark

We identified 13 major CAR-T clinical and commercial outcomes between 2017 and 2022. For each outcome, we identified the institutions and senior investigators associated with the research behind it. Sources included FDA approval records, company announcements, and securities filings.

The 13 outcomes include six FDA-approved CAR-T products, two acquisitions valued above $1 billion, and five major company foundings or public-market events.

Company foundings and public-market events (5). The remaining outcomes were associated with Allogene Therapeutics, Autolus Therapeutics, Precision BioSciences, Poseida Therapeutics, and Legend Biotech [31–35].

The two acquisitions alone had a combined disclosed value of $20.9 billion.

We defined this set of outcomes before comparing it with the framework ranking. This prevents us from choosing successful laboratories simply because the framework ranked them highly.

### 6.2 Identifying the Highest-Priority Papers and Laboratories

We used the framework to rank CAR-T research papers published in 2015 and 2016 according to their predicted probability of a paper-trial connection. The publication dataset included some papers from adjacent areas of cancer immunotherapy that were connected to CAR-T research but were not themselves primarily focused on CAR-T. For the industry-ranking analysis, we wanted to evaluate papers that a pharmaceutical scout would reasonably consider part of the CAR-T field. We therefore restricted the ranking to papers classified as CAR-T-related using OpenAlex topic information [18]. This produced a final ranking set of 488 papers.

We then examined the top-ranked papers and identified the laboratories responsible for them. A paper was counted as a hit if its laboratory was associated with at least one of the 13 predefined clinical or commercial outcomes.

This produces a direct measure of how concentrated the framework’s highest-ranked papers were among research groups associated with major CAR-T developments.

### 6.3 Evaluating the Ranking

Among the framework’s top 20 papers, 80% came from laboratories associated with one of the 13 predefined CAR-T clinical or commercial outcomes. Randomly selecting papers from the same pool produced a hit rate of 30% (Table 4).

**Table 3.** Predefined CAR-T clinical and commercial outcomes used as the retrospective benchmark.

| Outcome | Type | Year | Reference |
| --- | --- | --- | --- |
| Kymriah (tisagenlecleucel) | FDA approval | 2017 | [10] |
| Yescarta (axicabtagene ciloleucel) | FDA approval | 2017 | [24] |
| Tecartus (brexucabtagene autoleucel) | FDA approval | 2020 | [25] |
| Breyanzi (lisocabtagene maraleucel) | FDA approval | 2021 | [26] |
| Abecma (idecabtagene vicleucel) | FDA approval | 2021 | [27] |
| Carvykti (ciltacabtagene autoleucel) | FDA approval | 2022 | [28] |
| Kite Pharma acquisition by Gilead Sciences | Acquisition | 2017 | [29] |
| Juno Therapeutics acquisition by Celgene | Acquisition | 2018 | [30] |
| Allogene Therapeutics | Company/public-market event | 2018 | [31] |
| Autolus Therapeutics | Company/public-market event | 2018 | [32] |
| Precision BioSciences | Company/public-market event | 2019 | [33] |
| Poseida Therapeutics | Company/public-market event | 2020 | [34] |
| Legend Biotech | Company/public-market event | 2020 | [35] |

**Table 4.** Top-20 hit rate.

| Ranking | Top-20 hit rate | Outcomes represented |
| --- | --- | --- |
| Model ranking | 80.0% | 9 of 13 |
| Random | 30.0% | 5 of 13 |

The top 20 papers pointed to laboratories associated with nine of the 13 outcomes. These included the research groups connected with Kymriah, Yescarta, Tecartus, Breyanzi, Abecma, and Carvykti, as well as the acquisitions of Kite Pharma and Juno Therapeutics and the Legend Biotech public-market event.

The four outcomes not represented in the top 20 were associated with Allogene Therapeutics, Autolus Therapeutics, Precision BioSciences, and Poseida Therapeutics. A paper connected to the UCL and Cellectis research lineage appeared at rank 39, placing that research group outside the top 20 but still relatively high in the ranking.

These results suggest that the framework could have been useful as an early screening tool for pharmaceutical scouting. Rather than reviewing all CAR-T papers and laboratories with equal intensity, a company using the ranking could have concentrated its attention on a much smaller group containing a substantially higher proportion of research teams associated with important clinical and commercial developments.

## 7 Testing the Framework in a Second Therapeutic Area

The CAR-T analysis shows that paper-inherent information can strongly predict whether a research paper will be connected to a subsequent clinical trial within one therapeutic area. The next question is whether the same framework remains useful outside CAR-T.

We tested the framework in a second therapeutic area: checkpoint inhibitors. Checkpoint inhibitors are a different form of cancer immunotherapy from CAR-T and include therapies targeting immune-regulatory proteins such as PD-1, PD-L1, and CTLA-4. Using the same clinical-trial selection criteria applied to CAR-T, we identified 3,824 checkpoint-inhibitor trials from ClinicalTrials.gov [15]. Among papers published in 2015 and 2016, we identified 429 positive examples through direct trial references and sampled 900 negative examples, which were cross-checked against PubMed Knowledge Graph 2.0 [17].

We evaluated the framework in two ways.

First, we applied the CAR-T-trained model directly to the checkpoint-inhibitor papers without retraining it. This tests whether the relationships learned in CAR-T transfer to a different therapeutic area. The models achieved an AUC of 0.728.

Second, we trained a new model using the checkpoint-inhibitor data and evaluated it on a held-out 20% test set. This tests whether the same modeling framework remains predictive when the relationships between the indicators and clinical-trial connection are learned specifically for checkpoint inhibitors. The checkpoint-inhibitor model achieved an AUC of 0.777 (Figure 12).

**Fig. 12.**
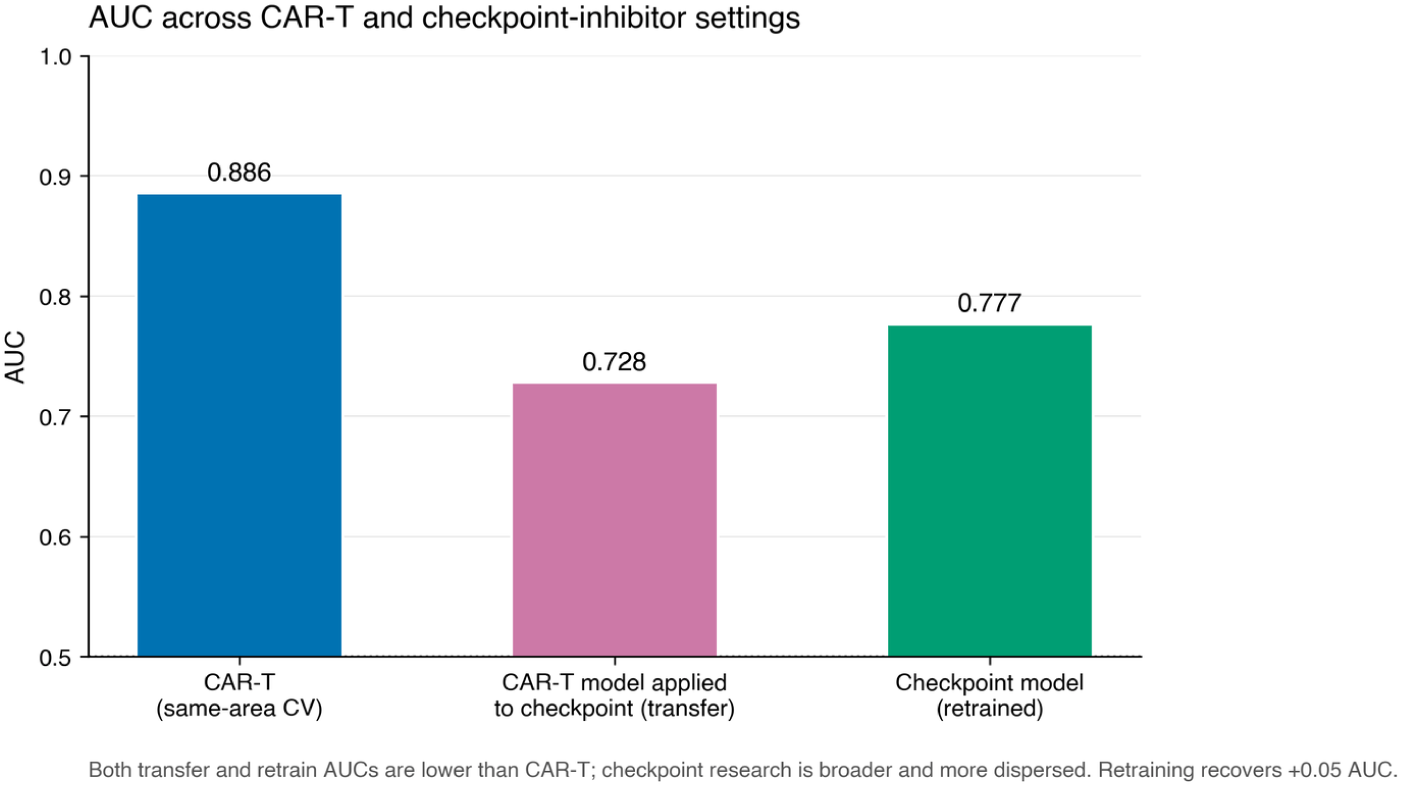
Cross-validated AUC in CAR-T alongside the transfer and retrained results for checkpoint inhibitors.

The difference between these two results is informative. An AUC of 0.728 without retraining indicates that some of the predictive relationships identified in CAR-T also appear in checkpoint-inhibitor research. The improvement to 0.777 after retraining indicates that other relationships are specific to the therapeutic area.

Performance in both tests was lower than in CAR-T. This suggests that the framework can extend beyond CAR-T, but that predictive performance may depend substantially on the structure of the therapeutic field being studied.

CAR-T may be particularly favorable for this type of prediction because its development was relatively concentrated across a smaller number of research groups and a more focused set of therapeutic programs. Checkpoint-inhibitor research spans a broader range of investigators, diseases, biological targets, and treatment settings. That greater heterogeneity may make clinical-trial connections more difficult to predict from publication characteristics alone.

Taken together, these results suggest that the framework is not limited to CAR-T, but that models are likely to benefit from being trained within individual therapeutic areas.

## 8 Discussion

This study suggests that the early identification of biomedical research with clinical potential may be more feasible than is commonly assumed.

Most systems for judging scientific promise rely heavily on signals that appear after publication: citations, journal visibility, reputation, funding, or expert attention. These signals can be useful, but they arrive only after the research has already begun to attract notice.

The results here point to a different possibility. A meaningful part of the signal appears to be present in the research paper itself. That means the decision problem may not always be “wait and see which papers attract attention,” but rather “identify useful patterns in the research before that attention develops.”

This has several broader implications.

### 8.1 Earlier identification of promising research

If strong predictions can be made from the paper itself, research organizations may be able to identify promising work earlier than they do today.

For pharmaceutical companies, this could change how scientific scouting is organized. Instead of beginning with highly cited papers, famous investigators, or well-known institutions, a screening system could first examine a much larger body of newly published work and identify a smaller set for expert review.

The same idea could apply inside universities, research institutes, and biotechnology companies. Large organizations often have more research activity than any individual decision-maker can evaluate closely. A model could help prioritize which papers, laboratories, or programs deserve additional attention.

The value of such a system would not be to replace scientific judgment. Its value would be to make that judgment more targeted.

### 8.2 A different way to think about scientific signals

The findings also raise a broader question about how research quality and clinical potential are recognized.

Citations and reputation are partly social signals. They reflect how other researchers respond to a paper and how the scientific community values the people and institutions behind it.

By contrast, the paper-inherent indicators describe the research itself: what is being studied, how the problem is framed, what language is used, and how the work is organized.

The strong performance of these indicators suggests that later clinical development may leave detectable traces in the scientific work before external recognition accumulates, consistent with what Nelson et al. [12] report for patent, guideline, and policy inclusion.

This does not mean that citations, reputation, or institutional resources are unimportant. It means that they may not be the only place to look for early evidence of which papers are likely to be connected to subsequent clinical trials.

## 9 Limitations

This study is a proof of concept, and several limitations affect how broadly the results should be interpreted.

First, the primary analysis is based on one therapeutic area. CAR-T has a relatively concentrated history, with a limited number of prominent laboratories and a clear sequence from research papers to clinical trials and commercial products. The lower performance in checkpoint inhibitors suggests that CAR-T may be an unusually favorable setting for this type of prediction. More therapeutic areas are needed before drawing broader conclusions.

Second, the labels are reconstructed rather than directly observed. ClinicalTrials.gov does not consistently record the research papers that preceded a trial, so many connections had to be recovered through references and citation chains. This means some true paper-to-trial connections were almost certainly missed, while some indirect connections may have been included even when the earlier paper played only a minor role. Relatedly, the pipeline does not perfectly distinguish preclinical papers from earlier trial reports, so some positives may themselves be reports of earlier trials being connected to clinical trials rather than preclinical work predicting a first clinical entry. Because the models are trained on these labels, any errors in label construction can affect all of the reported results.

Third, the retrospective laboratory test is not fully independent of the main analysis. Laboratories that later produced major CAR-T products or companies also tended to be prominent laboratories within the field. Prominence can affect publishing and citation patterns for reasons unrelated to whether any particular paper was connected to a clinical trial. The retrospective test therefore provides additional evidence, but it should not be treated as a completely independent validation.

Fourth, the class balance in the study does not represent the real-world frequency of success. The final dataset contains 453 positive papers and 897 negative papers, which creates a much higher proportion of positive examples than a pharmaceutical scout would encounter when screening biomedical research more broadly. AUC measures how well the model separates the two groups, but it does not tell us how many highly ranked papers would actually be connected to clinical trials in a real deployment setting; precision-recall analysis is more informative than ROC analysis under this kind of class imbalance [22].

Fifth, the outcome is broad. A paper is labeled positive if it is connected to a clinical trial, regardless of what happened afterward. A paper connected to an early Phase I safety study is therefore treated the same as one connected to a program that later reached approval. The model predicts whether a paper will be connected to a clinical trial, not whether the treatment will ultimately succeed.

## 10 Conclusion

This study shows that it is possible to predict which newly published biomedical research papers will be connected to subsequent clinical trials. Using CAR-T cell therapy as a proof-of-concept setting, we reconstructed connections between research papers and subsequent clinical trials and trained models to distinguish papers that were connected to subsequent clinical trials from those that were not.

The results show that this prediction can be made with high accuracy using information contained in the paper itself. Paper-inherent indicators describing scientific topic, language, biomedical orientation, and research-team structure achieved a cross-validated AUC of 0.908. First-year citations were also predictive, with an AUC of 0.797, while adding measures of author, journal, and institutional provenance increased the paper-inherent model only slightly, from 0.908 to 0.912. This indicates that much of the predictive information is already present in the research paper at the time of publication.

The model also produced a ranking with practical relevance. Among the top 20 CAR-T papers, 80% came from laboratories associated with one of 13 predefined major clinical or commercial outcomes. This suggests that the predictions could be useful not only for identifying individual papers with a greater likelihood of being connected to subsequent clinical trials, but also for helping pharmaceutical companies and research organizations identify laboratories and research programs that warrant closer evaluation.

Taken together, the findings show that which biomedical research papers will be connected to subsequent clinical trials can be predicted from characteristics observable at publication. The citation and provenance analyses provide additional evidence about the information available for this prediction, but the central result is that meaningful differences between papers that are and are not connected to subsequent clinical trials can be detected from newly published papers.

The broader implication is that biomedical research could be screened for translational potential earlier in the research lifecycle. With validation across additional therapeutic areas, this approach could provide a useful tool for prioritizing papers, laboratories, and therapeutic programs for closer scientific and commercial evaluation.

## Data Availability

The labeled dataset and the derived indicators supporting the findings of this study are available at https://github.com/jtbrennan/predicting-clinical-translation-of-biomedical-research. The underlying sources are publicly accessible: ClinicalTrials.gov, PubMed, PubMed Knowledge Graph 2.0, and OpenAlex.

## Supplementary information

Not applicable.

## Declarations

- **Funding:** The authors received no specific funding for this work.
- **Conflict of interest:** The authors declare no competing interests.
- **Ethics approval and consent to participate:** Not applicable. This study uses only publicly available bibliographic and clinical-trial registry data.
- **Consent for publication:** Not applicable.
- **Data availability:** The labeled dataset and the derived indicators supporting the findings of this study are available at https://github.com/jtbrennan/predicting-clinical-translation-of-biomedical-research. The underlying sources are publicly accessible: ClinicalTrials.gov [15], PubMed, PubMed Knowledge Graph 2.0 [17], and OpenAlex [18].
- **Materials availability:** Not applicable.
- **Code availability:** All code used to construct the dataset, compute the indicators, train the models and generate the figures is available at https://github.com/jtbrennan/predicting-clinical-translation-of-biomedical-research.
- **Author contribution:** J.B. conceived the study, designed the linking pipeline and indicator set, performed the analysis, produced the figures, and wrote the manuscript. J.S. and Y.Y. supervised the work and contributed to the study design, interpretation of results, and revision of the manuscript. All authors read and approved the final manuscript.

## Notes

### Competing Interest Statement

The authors have declared no competing interest.

## References

[1] Contopoulos-Ioannidis, D.G., Ntzani, E., Ioannidis, J.P.A.: Translation of highly promising basic science research into clinical applications. The American Journal of Medicine 114(6), 477–484 (2003) 10.1016/S0002-9343(03)00013-5

[2] Hackam, D.G., Redelmeier, D.A.: Translation of research evidence from animals to humans. JAMA 296(14), 1731–1732 (2006) 10.1001/jama.296.14.1731

[3] Lalu, M.M., Montroy, J., Begley, C.G., Bubela, T., Hunniford, V., Speck, M., Avey, M.T., Fergusson, D.A.: Identifying and understanding factors that affect the translation of therapies from the laboratory to patients: A study protocol. BMJ Open 10(9), 039699 (2020) 10.1136/bmjopen-2020-039699

[4] Sertkaya, A., Wong, H.-H., Jessup, A., Beleche, T.: Key cost drivers of pharmaceutical clinical trials in the United States. Clinical Trials 13(2), 117–126 (2016) 10.1177/1740774515625964

[5] Paul, S.M., Mytelka, D.S., Dunwiddie, C.T., Persinger, C.C., Munos, B.H., Lindborg, S.R., Schacht, A.L.: How to improve R&D productivity: The pharmaceutical industry’s grand challenge. Nature Reviews Drug Discovery 9(3), 203–214 (2010) 10.1038/nrd3078

[6] Peck, R.W., Lendrem, D.W., Grant, I., Lendrem, B.C., Isaacs, J.D.: Why is it hard to terminate failing projects in pharmaceutical R&D? Nature Reviews Drug Discovery 14(10), 663–664 (2015) 10.1038/nrd4725

[7] Weber, G.M.: Identifying translational science within the triangle of biomedicine. Journal of Translational Medicine 11, 126 (2013) 10.1186/1479-5876-11-126

[8] Hutchins, B.I., Davis, M.T., Meseroll, R.A., Santangelo, G.M.: Predicting translational progress in biomedical research. PLOS Biology 17(10), 3000416 (2019) 10.1371/journal.pbio.3000416

[9] Cappell, K.M., Kochenderfer, J.N.: Long-term outcomes following CAR T cell therapy: What we know so far. Nature Reviews Clinical Oncology 20(6), 359–371 (2023) 10.1038/s41571-023-00754-1

[10] U.S. Food and Drug Administration: KYMRIAH (tisagenlecleucel). https://www.fda.gov/vaccines-blood-biologics/cellular-gene-therapy-products/kymriah. BLA STN 125646. Initial approval 30 August 2017 (2017)

[11] Wang, D., Song, C., Barabási, A.-L.: Quantifying long-term scientific impact. Science 342(6154), 127–132 (2013) 10.1126/science.1237825

[12] Nelson, A.P.K., Gray, R.J., Ruffle, J.K., Watkins, H.C., Herron, D., Sorros, N., Mikhailov, D., Cardoso, M.J., Ourselin, S., McNally, N., Williams, B., Rees, G.E., Nachev, P.: Deep forecasting of translational impact in medical research. Patterns 3(5), 100483 (2022) 10.1016/j.patter.2022.100483

[13] Wang, S., Šuster, S., Baldwin, T., Verspoor, K.: Predicting publication of clinical trials using structured and unstructured data: Model development and validation study. Journal of Medical Internet Research 24(12), 38859 (2022) 10.2196/38859

[14] Fortunato, S., Bergstrom, C.T., Börner, K., Evans, J.A., Helbing, D., Milojević, S., Petersen, A.M., Radicchi, F., Sinatra, R., Uzzi, B., Vespignani, A., Waltman, L., Wang, D., Barabási, A.-L.: Science of science. Science 359(6379), 0185 (2018) 10.1126/science.aao0185

[15] U.S. National Library of Medicine: ClinicalTrials.gov. https://clinicaltrials.gov. National Institutes of Health. Accessed 2026 (2026)

[16] National Institutes of Health: Clinical trials: The basics. https://www.nih.gov/health-information/nih-clinical-research-trials-you/basics. U.S. Department of Health and Human Services. Accessed 2025 (2025)

[17] Xu, J., Yu, C., Xu, J., Torvik, V.I., Kang, J., Sung, M., Song, M., Bu, Y., Ding, Y.: PubMed knowledge graph 2.0: Connecting papers, patents, and clinical trials in biomedical science. Scientific Data 12(1), 1018 (2025) 10.1038/s41597-025-05343-8

[18] Priem, J., Piwowar, H., Orr, R.: OpenAlex: A fully-open index of scholarly works, authors, venues, institutions, and concepts. arXiv preprint arXiv:2205.01833 (2022) 10.48550/arXiv.2205.01833. Data accessed via the OpenAlex API, 2026

[19] U.S. National Library of Medicine: Medical Subject Headings (MeSH). https://www.nlm.nih.gov/mesh/. National Institutes of Health. Accessed 2026 (2026)

[20] Pedregosa, F., Varoquaux, G., Gramfort, A., Michel, V., Thirion, B., Grisel, O., Blondel, M., Prettenhofer, P., Weiss, R., Dubourg, V., Vanderplas, J., Passos, A., Cournapeau, D., Brucher, M., Perrot, M., Duchesnay, É.: Scikit-learn: Machine learning in Python. Journal of Machine Learning Research 12, 2825–2830 (2011)

[21] Nahm, F.S.: Receiver operating characteristic curve: Overview and practical use for clinicians. Korean Journal of Anesthesiology 75(1), 25–36 (2022) 10.4097/kja.21209

[22] Saito, T., Rehmsmeier, M.: The precision-recall plot is more informative than the ROC plot when evaluating binary classifiers on imbalanced datasets. PLOS ONE 10(3), 0118432 (2015) 10.1371/journal.pone.0118432

[23] Schuhmacher, A., Gassmann, O., Bieniok, D., Hinder, M., Hartl, D.: Open innovation: A paradigm shift in pharma R&D? Drug Discovery Today 27(9), 2395–2405 (2022) 10.1016/j.drudis.2022.05.018

[24] U.S. Food and Drug Administration: YESCARTA (axicabtagene ciloleucel). https://www.fda.gov/vaccines-blood-biologics/cellular-gene-therapy-products/yescarta-axicabtagene-ciloleucel. BLA STN 125643. Initial approval 18 October 2017 (2017)

[25] U.S. Food and Drug Administration: TECARTUS (brexucabtagene autoleucel). https://www.fda.gov/vaccines-blood-biologics/cellular-gene-therapy-products/tecartus. Initial approval 24 July 2020 (2020)

[26] U.S. Food and Drug Administration: BREYANZI (lisocabtagene maraleucel). https://www.fda.gov/vaccines-blood-biologics/cellular-gene-therapy-products/breyanzi. Initial approval 5 February 2021 (2021)

[27] U.S. Food and Drug Administration: ABECMA (idecabtagene vicleucel). https://www.fda.gov/vaccines-blood-biologics/abecma. BLA STN 125736. Initial approval 26 March 2021 (2021)

[28] U.S. Food and Drug Administration: CARVYKTI (ciltacabtagene autoleucel). https://www.fda.gov/vaccines-blood-biologics/cellular-gene-therapy-products/carvykti. Initial approval February 2022 (2022)

[29] Gilead Sciences: Gilead Sciences to acquire Kite Pharma for $11.9 billion. https://www.gilead.com/news/news-details/2017/gilead-sciences-to-acquire-kite-pharma-for-119-billion. Press release, 28 August 2017. Also filed as Exhibit 99.1 to Gilead Sciences Form 8-K, SEC CIK 0000882095 (2017)

[30] Celgene Corporation: Celgene to acquire Juno Therapeutics for $9.0 billion. https://www.sec.gov/Archives/edgar/data/0001594864/000119312518015255/d531432dex991.htm. Press release, 22 January 2018. Filed as Exhibit 99.1 to Juno Therapeutics Form 8-K, SEC CIK 0001594864 (2018)

[31] Allogene Therapeutics: Prospectus (initial public offering). https://www.sec.gov/Archives/edgar/data/1737287/000119312518297747/d548088d424b4.htm. SEC Form 424B4, filed 11 October 2018. Registration Nos. 333-227333 and 333-227774, CIK 0001737287 (2018)

[32] Autolus Therapeutics: Prospectus (initial public offering of American Depositary Shares). https://www.sec.gov/Archives/edgar/data/1730463/000119312518201011/d531832d424b4.htm. SEC Form 424B4, filed June 2018; IPO completed 22 June 2018. CIK 0001730463 (2018)

[33] Precision BioSciences: Registration statement (initial public offering). https://www.sec.gov/Archives/edgar/data/1357874/000119312519077399/d717633ds1a.htm. SEC Form S-1/A, filed 18 March 2019. Registration No. 333-230034, CIK 0001357874. IPO priced 27 March 2019 (2019)

[34] Poseida Therapeutics: Prospectus (initial public offering). https://www.sec.gov/Archives/edgar/data/1661460/000119312520191150/d872388d424b4.htm. SEC Form 424B4, filed 10 July 2020. Accession No. 0001193125-20-191150, File No. 333-239321, CIK 0001661460 (2020)

[35] Legend Biotech: Prospectus (initial public offering of American Depositary Shares). https://www.sec.gov/Archives/edgar/data/1801198/000119312520162919/d884493d424b4.htm. SEC Form 424B4, filed June 2020. Registration No. 333-238232, CIK 0001801198 (2020)

